# COVID-19 Vaccine Efficacy in a Diverse Urban Healthcare Worker Population

**DOI:** 10.1101/2021.09.02.21263038

**Authors:** Eirini Iliaki, Fan-Yun Lan, Costas A. Christophi, Guido Guidotti, Alexander D. Jobrack, Jane Buley, Neetha Nathan, Rebecca Osgood, Lou Ann Bruno-Murtha, Stefanos N. Kales

## Abstract

**Objective:** To investigate COVID-19 vaccine efficacy (VE) among healthcare workers (HCWs) in an ethnically diverse community healthcare system, during its initial immunization campaign.

**Methods:** HCWs of the system were retrospectively included from the beginning of a COVID-19 vaccination program (December 16, 2020) until March 31, 2021. Those with a prior COVID-19 infection before December 15 were excluded. The Occupational Health department of the system ran a COVID-19 screening and testing referral program for workers, consistently throughout the study period. A master database had been established and updated comprising of the demographics, COVID-19 PCR assays, and vaccinations of each HCW. Andersen-Gill extension of the Cox models were built to estimate the VE of fully/partially vaccinated person-days at risk.

**Results:** Among the 4317 eligible HCWs, 3249 (75%) received any vaccination during the study period. Vaccinated HCWs were older, less likely to be Black/African American, Hispanic/Latino or identify as two or more races, and more likely to be medical providers. After adjusting for age, sex, race, and the statewide background incidence at the time of vaccination, we observed a VE of 80.2% (95% CI: 57.5–90.8%) for ≧14 days after the first dose of Pfizer/Moderna, and 95.5% (95% CI: 88.2-98.3%) among those fully vaccinated (i.e. ≧14 days after the second dose of Pfizer/Moderna or the single dose of J&J/Janssen).

**Conclusion:** COVID-19 vaccine effectiveness in the real world is promising, and these data in concert with culturally appropriate may decrease vaccine hesitancy.

## Introduction

Healthcare workers (HCWs) have been essential workers in the fight against the SARS-CoV-2 (the virus causing COVID-19) pandemic.^1^ Due to close contact with patients and co-workers, healthcare workers have been at risk for COVID-19, and some have suffered from serious illness.^2-6^ Apart from workplace exposures, other sociodemographic risk factors for COVID-19 infection include race/ethnicity and incidence in the community of the employee’s residence.^5,7^ Although universal masking and other personal protective equipment (PPE) offer some protection,^8,9^ COVID-19 vaccines show the most promise for preventing SARS-CoV-2 among HCWs.^10^ Vaccine efficacy (VE) in the clinical trials was approximately 95% for fully immunized persons with both the Pfizer-BioNTech and Moderna mRNA vaccines^11,12^ and 66% for the J&J/Janssen vaccine.^13^ Real world data on vaccine effectiveness in the US have emerged from healthcare populations in large academic hospital systems showing encouraging results, including a study by the CDC team in 8 US locations enrolled in a longitudinal program,^14^ and a 90% vaccine effectiveness in the Mayo Clinic system amongst others.^15-17^

However, specific data on vaccine effectiveness from urban hospital settings with diverse employee populations are lacking, and most studies have not controlled for demographics including race and background community COVID-19 incidence, which have been demonstrated to be COVID-19 risk factors among HCWs. Our demographically diverse workforce is representative of its surrounding community, and many of our employees reside in communities that were disproportionately affected by COVID-19.^18^

Therefore, we analyzed employee records at our socio-demographically diverse community healthcare system for COVID-19 vaccination and molecular-testing-confirmed cases. Our analyses accounted for prior COVID-19 infection and controlled for age, sex, race and background rate of COVID-19 in the state during the study period.

## Methods

### Study Population and Design

The active serving (as of March 31, 2021) HCWs of a public, community-based healthcare organization in the Greater Boston area of Massachusetts, USA older than 18 years of age were studied retrospectively. As described previously,^19^ the occupational health service of the organization implemented a staff “COVID-19 hotline” system on March 9, 2020 for HCW-COVID-19-related concerns; electronically documented clinical information; and coordinated testing, result communication and return to work.

Employees were offered same day scheduling of a molecular test for SARS-CoV-2, for possible viral symptoms, voluntary asymptomatic screening for COVID-19 and advice on testing and quarantine in cases of potential contact with a COVID-19-positive/suspect person or travel. A standard triage form summarized elsewhere^19^ was completed by occupational health personnel based on contemporaneous telephonic interviews with each HCW. The form captures demographics, administrative information, potential exposure history, and eleven potential COVID-19 related viral symptoms.^19^

In addition, the Occupational Health department coordinated periodic asymptomatic voluntary screening testing for those employees working with high risk-populations, including geriatrics, inpatient psychiatry, and those visiting congregate care settings. Mandatory weekly screenings of employees visiting/working in long-term care facilities and home healthcare settings began on July 2, 2020 and included 107 employees.

Masking was obligatory since March 26, 2020, and patient-facing staff wore eye protection in addition to appropriate medical masking, as well as gowns for contact with known COVID-19 positive patients. Common spaces and break rooms were adjusted to accommodate for social distancing. Public mask mandates were in effect in all of Massachusetts during the present study period.

From December 15, 2020 to March 31, 2021 we conducted a retrospective review of all HCW with newly diagnosed COVID-19. The data we used were de-identified, the need for a consent was waived and the study was exempted by the Cambridge Health Alliance Institutional Review Board. (Protocol number 4/29/202-003)

### Vaccination Program and data collection

The Pfizer and Moderna vaccines were made available to HCWs starting on December 16 and December 23, 2020, respectively. Virtual town hall meetings were held by hospital leadership to educate employees. Massachusetts Department of Public Health state guidelines highlighted a community approach and prioritized vaccinating first all clinical and non-clinical staff working together regardless of their direct roles in high-risk areas. On December 29, 2020, two weeks after the vaccination program began, availability was opened to all hospital employees, including those with no direct patient contact and working remotely. Participation was voluntary by walking into our conveniently located hospital based vaccination sites and no appointment was required. After receiving its Emergency Use Authorization (EUA) in February 2021, a limited number of J&J/Janssen COVID-19 vaccine doses were included in the vaccination program. To improve initial vaccine acceptance, an additional town hall was presented on February 28, 2021 by a multi-racial and multi-ethnic panel to further promote vaccine information. A warning about the risk of vaccine induced thrombotic thrombocytopenia from the J&J/Janssen was communicated to our employees on April 13, 2021. Moreover, several additional focus-group discussions in different languages were held to further increase vaccine uptake.

Occupational health vaccine administration records were matched against a human resources roster of active employees of the healthcare organization to create a unified database including demographics, vaccination details, and SARS-CoV2 infection history.

### Vaccine Breakthrough Infections

We obtained PCR cycle threshold (CT) values on all positive samples run on the Hologic Panther Fusion system utilizing the Aptima SARS-CoV-2 PCR assay from immunized HCWs, and the samples were sent to the Commonwealth of Massachusetts laboratory for whole genome sequencing to potentially identify variants of concern after approval by the State epidemiologist. Employees with breakthrough infections had follow up by our Occupational health medical staff via telephonic interviews following the same protocol that we use for COVID-19 infected employees which includes recording dates of symptoms, potential exposures, types and severity of symptoms, employee risk factors, vaccine dates and type as well as contact investigation when indicated.

### Statistical Analysis

HCWs with a documented history of COVID-19 infection prior to December 15, 2020 were excluded from the main analyses. First, we compared the characteristics between the HCWs having received any vaccination by March 31, 2021 versus those that did not, using the chi-square test of independence for categorical variables, or the t-test for continuous variables, after checking for normality of the distribution.

Next, we analyzed VE as a function of SARS-CoV-2 infection rates calculated as incident cases per person-days at risk as follows. All HCWs’ person-days at risk were categorized based on time in each distinct vaccination status: 1) unvaccinated person-days, 2) person-days within 14 days after the 1^st^ dose (except for those receiving J&J/Janssen), 3) person-days over 14 days after the 1^st^ dose and prior to the 2^nd^ dose (except for those receiving J&J/Janssen), 4) person-days within 14 days after the 2^nd^ dose; or person-days within 14 days after the single dose (for those receiving J&J/Janssen), and 5) “fully-vaccinated” - person-days over 14 days after the 2^nd^ dose; or person-days over 14 days after the single dose (for those receiving J&J/Janssen).

For each category, we counted the number of COVID-19 infections and calculated the crude incidence rate per 10,000 person-days. Kaplan-Meier curves were used to visualize person-days of infection-free survival across the exposure categories. Since a HCW could contribute to person-days in multiple categories, we built an Andersen-Gill extension of the Cox proportional hazards models to account for correlated data,^14^ and further adjusted for potential confounders including age, sex, race, and the Massachusetts statewide 7-day average of new cases^20^ on the date of the first vaccine dose. The VE was calculated as 100% × (1-hazard ratio), and 95% confidence intervals (95% CIs) were presented along with the point estimates. Statistical analyses were performed using the SAS software version 9.4 (SAS Inc, Cary, NC, USA) and the R software (version 3.6.3). All P values presented are two-tailed and a P <.05 was considered statistically significant.

## Results

Among the 4635 healthcare system employees, before and during the study period, none of the 318 HCWs with a history of COVID-19 infection prior to December 15, 2020 experienced reinfection regardless of COVID-19 immunization status.

After excluding the above group with prior infection, a total of 4317 HCWs were included in the main analyses. Among them, 3249 (75%) had received any vaccination by the end of the study period (March 31, 2021) (Table 1). HCWs who had any immunization were older (45.7 ± 13.5 years old vs. 41.3 ± 12.8 years old, P<.001), more likely to be non-Hispanic/Latino whites. In addition, medical providers were more likely to be vaccinated compared with other HCWs (89% vs. 73%, P<.001) (Table 1). No significant adverse reactions of vaccination were reported among our population.

**Table 1.**
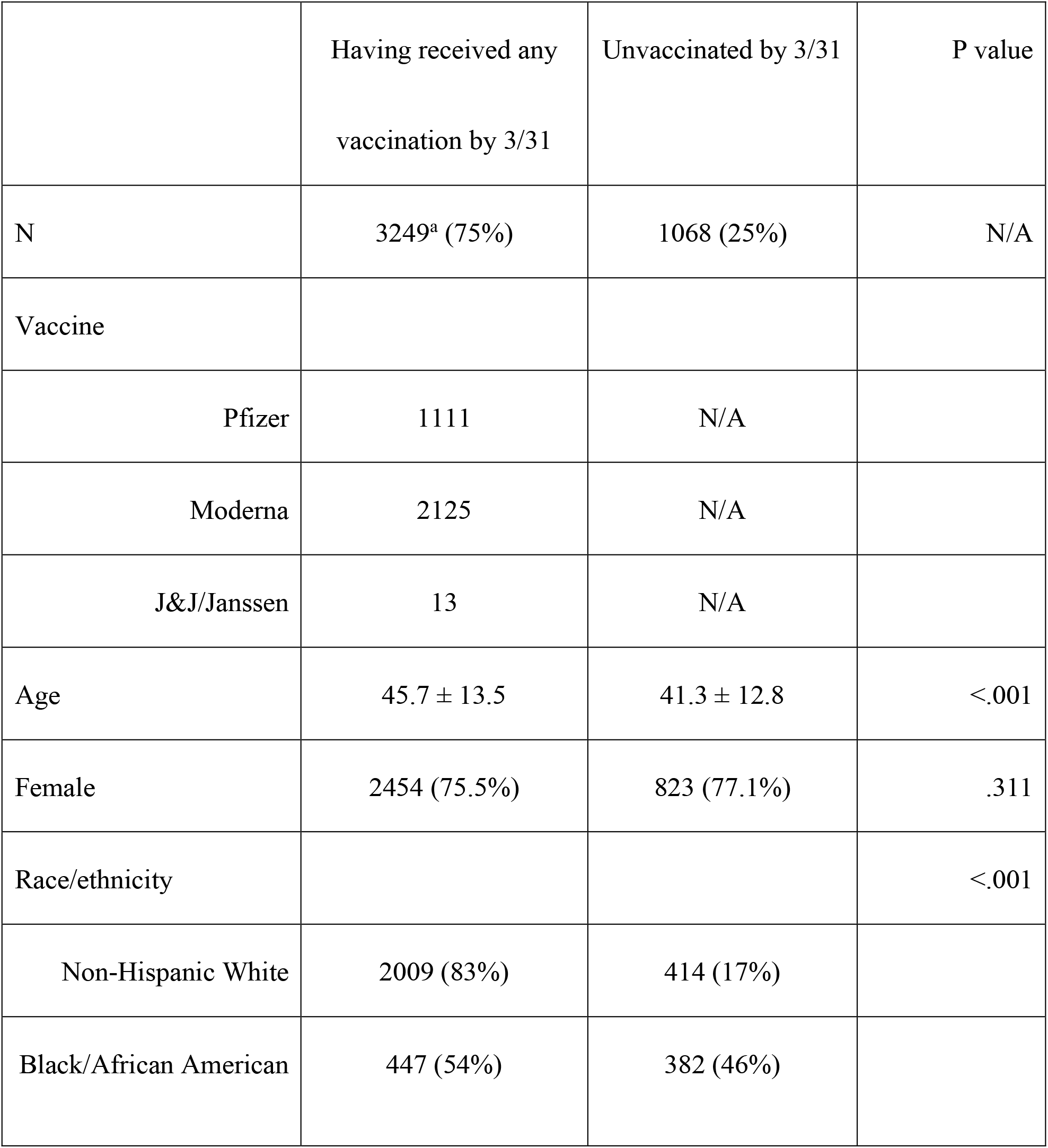
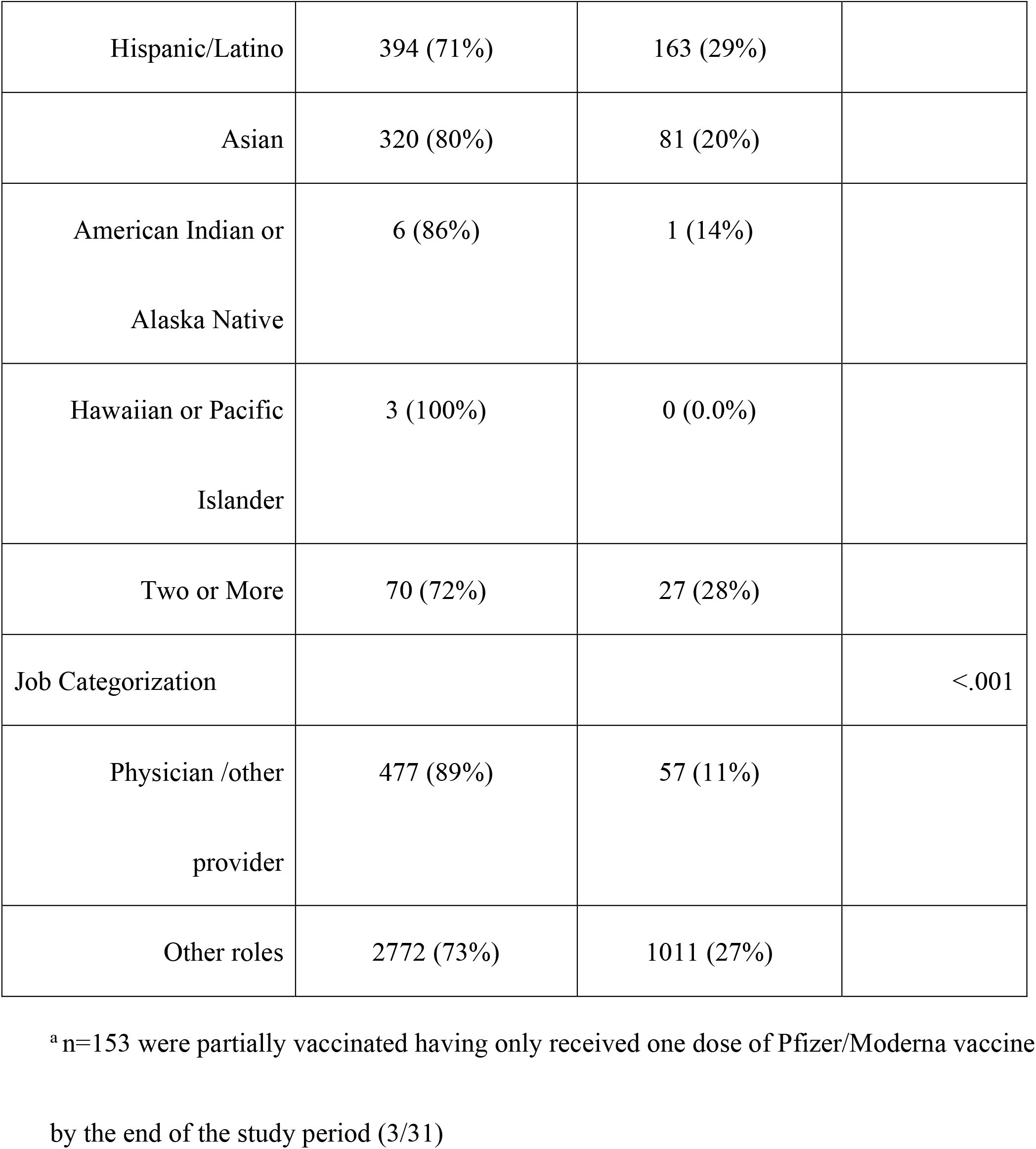
Characteristics comparing the healthcare workers receiving at least one vaccination by Mar 31, 2021 versus those remained unvaccinated (excluding 318 people infected before Dec 15, 2020)

|  | Having received any<br>vaccination by 3/31 | Unvaccinated by 3/31 | P value |
| --- | --- | --- | --- |
| N | 3249<sup>a</sup> (75%) | 1068 (25%) | N/A |
| Vaccine |  |  |  |
| Pfizer | 1111 | N/A |  |
| Moderna | 2125 | N/A |  |
| J&J/Janssen | 13 | N/A |  |
| Age | 45.7 ± 13.5 | 41.3 ± 12.8 | <.001 |
| Female | 2454 (75.5%) | 823 (77.1%) | .311 |
| Race/ethnicity |  |  | <.001 |
| Non-Hispanic White | 2009 (83%) | 414 (17%) |  |
| Black/African American | 447 (54%) | 382 (46%) |  |
| Hispanic/Latino | 394 (71%) | 163 (29%) |  |
| Asian | 320 (80%) | 81 (20%) |  |
| American Indian or<br>Alaska Native | 6 (86%) | 1 (14%) |  |
| Hawaiian or Pacific<br>Islander | 3 (100%) | 0 (0.0%) |  |
| Two or More | 70 (72%) | 27 (28%) |  |
| Job Categorization |  |  | <.001 |
| Physician /other<br>provider | 477 (89%) | 57 (11%) |  |
| Other roles | 2772 (73%) | 1011 (27%) |  |

<sup>a</sup> n=153 were partially vaccinated having only received one dose of Pfizer/Moderna vaccine by the end of the study period (3/31)

The 4317 HCWs contributed to a total of 444,058 person-days at risk (Table 2). The crude infection rates across the five categories of vaccination status were 7.69, 6.94, 1.97, 0.48, and 0.27, respectively (Table 2, Figure 1). During the study period, the statewide 7-day averaged incidence rate fell from 3,257 cases per day on the date of start to the to 1,563 at the end of the study.

**Table 2.** Rate of infection during the study period across the five vaccination categories (separating period with first dose only to <14 and 14+ days and excluding 318 people infected before Dec 15, 2020)

**infected before Dec 15, 2020)**
| Status | Person-days | No of infections | Rate per 10,000 person-days | Unadjusted vaccine effectiveness % (95% CI) | Adjusted vaccine effectiveness % (95% CI)<sup>a</sup> |
| --- | --- | --- | --- | --- | --- |
| Unvaccinated | 172845 | 133 | 7.69 | NA | NA |
| First dose (<14 days) | 40344 | 28 | 6.94 | 26.9<br>(-17.6–54.5) | 28.1<br>(-15.9–55.4) |
| First dose (14+ days) | 40577 | 8 | 1.97 | 80.1<br>(57.8–90.6) | 80.2<br>(57.5–90.8) |
| Second dose | 41817 | 2 | 0.48 | 95.4<br>(80.8–98.9) | 95.2<br>(80.0–98.8) |
| Fully vaccinated | 148475 | 4 | 0.27 | 97.2<br>(92.5–99.0) | 95.5<br>(88.2–98.3) |

Vaccine effectiveness (95% CI) derived from the Andersen-Gill extension of the Cox proportional hazards model
<sup>a</sup> Adjust for age, sex, race, and the Massachusetts statewide 7-day average of new cases at the date for the first vaccine dose. Those with the race of “American Indian or Alaska Native”, “Hawaiian or Pacific Islander”, or “Two or More” were pooled into one level “other race”.

**Figure 1.**
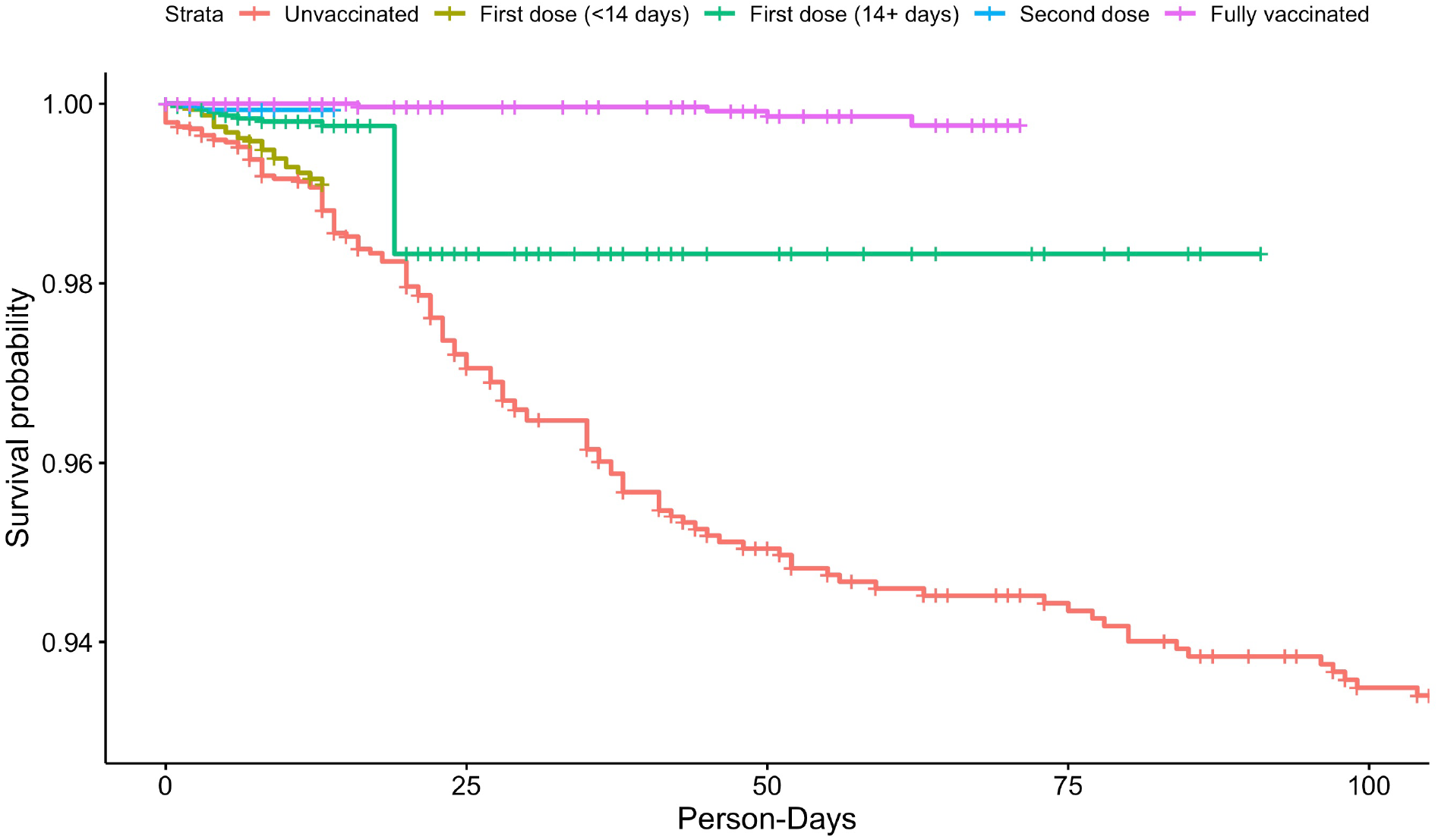
The Kaplan-Meier curve for the survival (i.e. infection-free) person-days across the five categories based on vaccination status.

Using the Andersen-Gill extension of the Cox proportional hazards model (adjusted for age, sex, race, and the Massachusetts statewide 7-day average of new cases), we observed an effectiveness of 80.2% at 14 or more days after the first dose of Pfizer/Moderna. A receiving two doses of Pfizer/Moderna or single dose of J&J/Janssen adjusted effectiveness increased to 95.2% (95% CI: 80.0-98.8), and 95.5% (95% CI: 88.2-98.3) among those fully vaccinated (Table 2 and Figure 1).

### Vaccine Breakthrough Infections

During the study period, there were six breakthrough infections among fully vaccinated personnel. None of these employees required hospitalization and symptoms were mild for five and moderate for one. All but one were due to household exposure to family members. One had an immunocompromising condition. Two of the six had a CT value above 30. The state lab was not able to isolate any viral material for one of the six, raising suspicion of a false positive. None of the other five samples sent for genomic sequencing revealed any variants of concern.

## Discussion

Our study suggests a very high, pre-Delta variant vaccine effectiveness in the HCW population of a university-affiliated urban healthcare system, with an estimated adjusted VE for fully vaccinated individuals 95.5% (95% CI: 88.2–98.3%). The vast majority received mRNA vaccines (only 13 out of 3461 employees opted to receive J&J/Janssen). Our study results add to the emerging literature of the robust “real-world” VE among fully vaccinated HCW and are similar to the studies conducted in US healthcare populations. A CDC review of the literature from the US within a similar time frame shows five studies involving HCW vaccination;^21^ of the four that calculated VE 14 days after the second vaccine dose of the mRNA vaccines, VE ranged from 87-99%.^14,17,22,23^ Beyond the US, similar results have been seen in HCWs in vaccine effectiveness studies from the UK, Israel, Italy, and Denmark and range from 86%-97%, all measuring at least 7 days after the second dose.^24-30^ Our study is one of few controlling for demographics including race and background community COVID-19 incidence.

Real world data from the broader US population also show encouraging results as there were only 10,262 breakthrough infections recorded until April 30, 2021 among 101 million fully vaccinated Americans.^31^ Israel has had similar experience with surveillance data from Jan 23 to April 3, 2021, showing a 95.3% vaccine effectiveness and notably 97.2% (95% CI: 96.8–97.5%) decrease in hospitalizations and 96.7% (95% CI: 96.0–97.3%) in COVID-19 related deaths.^32^

It is worth noting that we observed that in the two weeks after the first vaccination the adjusted VE is very small (28%), likely reflecting the time period needed to build immune response. Although VE in our data set increases 14 days after the first dose to 80.2%, which is mirrored in the findings of other US HCW studies,^14,17,22^ it is important to highlight that the second vaccine dose seems essential to increase the strength and likely the durability of the immune response. This is supported by data on the new delta variant from the UK that clearly show that there is a reduction in VE of 15-20% after a receipt of only a single vaccine dose.^33^

Our study was conducted in an ethnically diverse population (non-white employees represented 44% of our employee population), where vaccination uptake for employees identifying as Black/African American was only 54%, which is much lower than white non-Hispanic employees (83%). This large difference in vaccine hesitancy/acceptance is a serious issue that merits further study and intervention. Out of the five studies in healthcare workers in the US within similar study period,^21^ only 3 report adequate data on employee race.^14,17,34^ HEROES-RECOVER is a network of longitudinal cohorts in eight locations (Phoenix, Tucson, and other areas in Arizona; Miami, Florida; Duluth, Minnesota; Portland, Oregon; Temple, Texas; and Salt Lake City, Utah) that includes healthcare personnel, first-responders, and other essential frontline workers. They report a higher vaccine acceptance on employees identifying as non-White for race than ours (68%) but only a small proportion of participants (14%) were non-white,^14^ revealing likely participation and sampling bias. The second study is the one that includes HCWs at the Mayo Clinic medical system with employees in Minnesota, Florida and Arizona, where employees who identified as black were only 4% of the 71,152 workforce.^17^ Vaccine acceptance among blacks was 47%, which is similar to ours (54%),^17^ as it was at St Jude’s Children’s hospital (Tennessee), where 22% of employees identify as black and their vaccine acceptance was 52%.^34^

On the other hand, our vaccine acceptance within the healthcare system was more than double that of citizens identifying as Black in the Commonwealth of MA-only 22% had received at least one COVID-19 vaccination as of March 30, 2021.^35^ The reasons for differential racial vaccine hesitancy/acceptance rates are most likely socioeconomic.^36^ This is consistent with our finding that clinical providers with generally higher socioeconomic status had higher vaccination rates (89%) when compared to non-provider HCWs (73%). These data support greater education and outreach efforts be focused on non-white and non-provider groups.

We had few breakthrough infections during the study period (albeit is was conducted prior to the arrival of delta in Massachusetts), with the majority (five out of six) cases being very mild and none resulting in hospitalizations or death. No variants of concern were discovered amongst the samples that were genotyped.

Our study has several strengths including that other than age and gender-we adjusted for race/ethnicity and 7-day incidence in MA at the time of vaccination to account for the dropping background rate of COVID-19 during the study period. Our population is multiethnic and 44% non-white which provides a great lens into communities with such makeup and allows us to draw better conclusions about such populations which are often under-represented in vaccine trials. It is worth noting that of the five studies in healthcare populations in the US, adequate race/ethnicity data are reported only in three. To study and address this phenomenon better it is imperative that data on race and ethnicity is gathered.

Limitations include the fact that this was not a randomised clinical trial and may be subject to biases and confounders that could skew the results. For example, vaccinated individuals may take more precautions due to being more health-conscious, but conversely may take fewer precautions due to perceived vaccination-induced protection. Thus, we do not expect any significant influence on our results. The infection rates decreased during the study period and may have potentiated the vaccine effectiveness, although we controlled for the 7-day MA infection rate. Since we do not conduct regular surveillance of the entire HCW population, there may have been additional asymptomatic breakthrough infections. Nonetheless, no asymptomatic cases were detected for the subset of HCW participating in surveillance testing, and there were no clusters of employee infections connected to non fully vaccinated individuals since the vaccination program started until the end of the study. This points to the fact that even if there was forward transmission, this was largely asymptomatic and non-consequential.

## Conclusion

Vaccination of HCWs is safest and effective and real-world protection during the study period among an ethnically diverse population was very similar (about 95%) to that achieved in clinical trials. We did observe a disparity with lower vaccine acceptance among non-white employees and those who are not medical providers. Efforts to develop culturally appropriate educational outreach to involve persons of different ethnicities and different healthcare roles should be explored to decrease vaccine hesitancy.

## Data Availability

De-identified data are available on reasonable request.

## Acknowledgements

The authors would like to thank their colleagues on CHA’s Vaccine Task Force for their efforts and leadership, as well as the employees of CHA for their service during the pandemic and overall high trust in the vaccine campaign.

## Abbreviations

HCW: healthcare worker
COVID-19: coronavirus disease 2019
VE: vaccine efficacy
CT: cycle threshold

## References

1. Bauchner H, Easley TJ, on behalf of the entire editorial and publishing staff of JAMA and the JAMA Network. Health care heroes of the COVID-19 pandemic. JAMA. 2020;323(20):2021.

2. Lan FY, Wei CF, Hsu YT, Christiani DC, Kales SN. Work-related COVID-19 transmission in six Asian countries/areas: A follow-up study. PLoS One. 2020;15(5):e0233588.

3. Kambhampati AK, O’Halloran AC, Whitaker M, et al. COVID-19-associated hospitalizations among health care personnel - COVID-NET, 13 States, March 1-May 31, 2020. MMWR Morb Mortal Wkly Rep. 2020;69(43):1576–1583.

4. CDC COVID-19 Response Team. Characteristics of health care personnel with COVID-19 - United States, February 12-April 9, 2020. MMWR Morb Mortal Wkly Rep. 2020;69(15):477–481.

5. Chou R, Dana T, Selph S, Totten AM, Buckley DI, Fu R. Update Alert 9: Epidemiology of and risk factors for coronavirus infection in health care workers. Ann Intern Med. 2021;174(7):W63–W64.

6. Chou R, Dana T, Buckley DI, Selph S, Fu R, Totten AM. Epidemiology of and risk factors for coronavirus infection in health care workers: A living rapid review. Ann Intern Med. 2020;173(2):120–136.

7. Jacob JT, Baker JM, Fridkin SK, et al. Risk factors associated with SARS-CoV-2 seropositivity among US health care personnel. JAMA Netw Open. 2021;4(3):e211283.

8. Lan FY, Christophi CA, Buley J, et al. Effects of universal masking on Massachusetts healthcare workers’ COVID-19 incidence. Occup Med (Lond). 2020;70(8):606–609.

9. Wang X, Ferro EG, Zhou G, Hashimoto D, Bhatt DL. Association between universal masking in a health care system and SARS-CoV-2 positivity among health care workers. JAMA. 2020;324(7):703–704.

10. Dooling K, McClung N, Chamberland M, et al. The advisory committee on immunization practices’ interim recommendation for allocating initial supplies of COVID-19 vaccine - United States, 2020. MMWR Morb Mortal Wkly Rep. 2020;69(49):1857–1859.

11. Polack FP, Thomas SJ, Kitchin N, et al. Safety and efficacy of the BNT162b2 mRNA Covid-19 vaccine. N Engl J Med. 2020;383(27):2603–2615.

12. Baden LR, El Sahly HM, Essink B, et al. Efficacy and safety of the mRNA-1273 SARS-CoV-2 vaccine. N Engl J Med. 2021;384(5):403–416.

13. Sadoff J, Gray G, Vandebosch A, et al. Safety and efficacy of single-dose Ad26.COV2.S vaccine against Covid-19. N Engl J Med. 2021;384(23):2187–2201.

14. Thompson MG, Burgess JL, Naleway AL, et al. Interim estimates of vaccine effectiveness of BNT162b2 and mRNA-1273 COVID-19 vaccines in preventing SARS-CoV-2 infection among health care personnel, first responders, and other essential and frontline workers - Eight U.S. locations, December 2020-March 2021. MMWR Morb Mortal Wkly Rep. 2021;70(13):495–500.

15. Keehner J, Horton LE, Pfeffer MA, et al. SARS-CoV-2 Infection after vaccination in health care workers in California. N Engl J Med. 2021;384(18):1774–1775.

16. Daniel W, Nivet M, Warner J, Podolsky DK. Early evidence of the effect of SARS-CoV-2 vaccine at one medical center. N Engl J Med. 2021;384(20):1962–1963.

17. Swift MD, Breeher LE, Tande AJ, et al. Effectiveness of mRNA COVID-19 vaccines against SARS-CoV-2 infection in a cohort of healthcare personnel [published online ahead of print Apr 26, 2021]. Clin Infect Dis. 2021;ciab361. https://doi.org/10.1093/cid/ciab361.

18. Lan FY, Filler R, Mathew S, et al. Sociodemographic risk factors for coronavirus disease 2019 (COVID-19) infection among Massachusetts healthcare workers: A retrospective cohort study [published online ahead of print Jan 28, 2021]. Infect Control Hosp Epidemiol. 2021;1–6. https://doi.org/10.1017/ice.2021.17.

19. Lan FY, Filler R, Mathew S, et al. COVID-19 symptoms predictive of healthcare workers’ SARS-CoV-2 PCR results. PLoS One. 2020;15(6):e0235460.

20. Massachusetts Department of Public Health. Archive of COVID-19 cases in Massachusetts. https://www.mass.gov/info-details/archive-of-covid-19-cases-in-massachusetts. Accessed August 23, 2021.

21. Centers for Disease Control and Prevention (CDC). Science Brief: COVID-19 Vaccines and Vaccination. https://www.cdc.gov/coronavirus/2019-ncov/science/science-briefs/fully-vaccinated-people.html. Accessed August 23, 2021.

22. Pilishvili T, Fleming-Dutra KE, Farrar JL, et al. Interim estimates of vaccine effectiveness of Pfizer-BioNTech and Moderna COVID-19 vaccines among health care personnel - 33 U.S. sites, January-March 2021. MMWR Morb Mortal Wkly Rep. 2021;70(20):753–758.

23. Cavanaugh AM, Fortier S, Lewis P, et al. COVID-19 outbreak associated with a SARS-CoV-2 R.1 lineage variant in a skilled nursing facility after vaccination program - Kentucky, March 2021. MMWR Morb Mortal Wkly Rep. 2021;70(17):639–643.

24. Lumley SF, Rodger G, Constantinides B, et al. An observational cohort study on the incidence of SARS-CoV-2 infection and B.1.1.7 variant infection in healthcare workers by antibody and vaccination status [published online ahead of print Jul 3, 2021]. Clin Infect Dis. 2021;ciab608. https://doi.org/10.1093/cid/ciab608.

25. Hall VJ, Foulkes S, Saei A, et al. COVID-19 vaccine coverage in health-care workers in England and effectiveness of BNT162b2 mRNA vaccine against infection (SIREN): a prospective, multicentre, cohort study. Lancet. 2021;397(10286):1725–1735.

26. V Shah As, Gribben C, Bishop J, et al. Effect of vaccination on transmission of COVID-19: an observational study in healthcare workers and their households. Preprint at https://www.medrxiv.org/content/10.1101/2021.03.11.21253275v1 (2021).

27. Angel Y, Spitzer A, Henig O, et al. Association between vaccination with BNT162b2 and incidence of symptomatic and asymptomatic SARS-CoV-2 infections among health care workers. JAMA. 2021;325(24):2457–2465.

28. Regev-Yochay G, Amit S, Bergwerk M, et al. Decreased infectivity following BNT162b2 vaccination: A prospective cohort study in Israel. Lancet Reg Health Eur. 2021;7:100150.

29. Fabiani M, Ramigni M, Gobbetto V, Mateo-Urdiales A, Pezzotti P, Piovesan C. Effectiveness of the Comirnaty (BNT162b2, BioNTech/Pfizer) vaccine in preventing SARS-CoV-2 infection among healthcare workers, Treviso province, Veneto region, Italy, 27 December 2020 to 24 March 2021. Euro Surveill. 2021;26(17):2100420.

30. Moustsen-Helms IR, Emborg H-D, Nielsen J, et al. Vaccine effectiveness after 1st and 2nd dose of the BNT162b2 mRNA Covid-19 Vaccine in long-term care facility residents and healthcare workers – a Danish cohort study. Preprint at https://www.medrxiv.org/content/10.1101/2021.03.08.21252200v1 (2021).

31. CDC COVID-19 Vaccine Breakthrough Case Investigations Team. COVID-19 vaccine breakthrough infections reported to CDC - United States, January 1-April 30, 2021. MMWR Morb Mortal Wkly Rep. 2021;70(21):792–793.

32. Haas EJ, Angulo FJ, McLaughlin JM, et al. Impact and effectiveness of mRNA BNT162b2 vaccine against SARS-CoV-2 infections and COVID-19 cases, hospitalisations, and deaths following a nationwide vaccination campaign in Israel: an observational study using national surveillance data. Lancet. 2021;397(10287):1819–1829.

33. Public Health England. 18 June 2021 Risk assessment for SARS-CoV-2 variant: Delta (VOC-21APR-02, B.1.617.2). https://assets.publishing.service.gov.uk/government/uploads/system/uploads/attachment_data/file/994761/18_June_2021_Risk_assessment_for_SARS-CoV-2_variant_DELTA.pdf. Accessed August 23, 2021.

34. Tang L, Hijano DR, Gaur AH, et al. Asymptomatic and symptomatic SARS-CoV-2 infections after BNT162b2 vaccination in a routinely screened workforce. JAMA. 2021;325(24):2500–2502.

35. Massachusetts Department of Public Health. Weekly COVID-19 Vaccination Report - Thursday, April 1, 2021. https://www.mass.gov/doc/weekly-covid-19-vaccination-report-april-1-2021/download. Accessed August 23, 2021.

36. Shekhar R, Sheikh AB, Upadhyay S, et al. COVID-19 vaccine acceptance among health care workers in the United States. Vaccines (Basel). 2021;9(2):119.

